# Vague retellings of personal narratives in temporal lobe epilepsy

**DOI:** 10.1101/2022.10.24.22281484

**Authors:** Fiore D’Aprano, Charles B. Malpas, Stefanie E. Roberts, Michael M. Saling

## Abstract

Aside from deficits identified in lexical retrieval, individuals with temporal lobe epilepsy (TLE) exhibit clinical oddities, such as circumstantiality in their language production. This becomes particularly evident when elicitation tasks impose minimal structure, or when impersonal narratives are retold over consecutive occasions. Personal reminiscence is highly specific and localised in time, placing specific demands on cognitive-linguistic systems. It is presumed that the nature of this elicitation paradigm will produce a unique psycholinguistic phenotype in those with TLE. Among controls there is a compression of output for impersonal narratives, but the opposite effect when personal narratives are retold. To investigate the micro- and macrolinguistic processes underpinning personal discourse production in TLE, we examined the elicited language output of in 15 surgically naïve individuals with TLE and 14 healthy controls. Participants were asked to recall and re-tell an autobiographical memory on four immediately consecutive occasions, representing an alternative unstructured elicitation. Following transcription and coding of output, a detailed multi-level discourse analysis of output volume, fluency, cohesion, and coherence was conducted. There were significant group by trial interactions for the number of novel units, the number of non-progression units, and for the proportion of non-progression to novel content. As anticipated, a distinctly different pattern emerged in TLE when compared with controls who did not compress their output volume across repetitions but instead produced greater novelty, and a more coherent and refined account over time. Individuals with TLE consistently told a less distinct story across repetitions, with disturbances in fluency, cohesion, and coherence. This reflects a reduced capacity to produce a coherent mental representation, in all likelihood related to the neurolinguistic demands of recalling and retelling specific personal events.

## Introduction

Circumstantial language in individuals with temporal lobe epilepsy (TLE) is clinically identifiable and is described as pedantic and repetitive (Bear et al., 1982; Benson, 1991; Geschwind, 1977). TLE is frequently associated with cognitive-linguistic impairments (Zhao et al., 2014) which, in current practice, are most commonly demonstrated on neuropsychological examination at a single-word level (Bartha et al., 2005; Dutta et al., 2018). At the level of discourse, individuals with TLE exhibit an impairment in cohesion and coherence when impersonal narratives based on an external visual stimulus (that is, a structured elicitation context) such as a cartoon, are retold (D’Aprano et al., 2022b pre-print; Field et al., 2000). Contexts that elicit spontaneous narrative are cognitively demanding because they rely on internal generation of ideas and message-level planning in contrast to those that provide external structure and prompting (Barch & Berenbaum, 1997). Individuals with TLE become more verbose, less concise, less informative, and less coherent than healthy controls when contextual structure is low (D’Aprano, Malpas, Roberts, & Saling, 2022a pre-print). In that study we found that their production of extraneous details, elaborations, and self-disclosures increased when asked to recall the way in which their usually daily activities unfold (Typical Day). We suggested that this phenomenon relates to greater freedom in lexical and propositional selection within a familiar though unrestricted semantic space (D’Aprano et al., 2022a pre-print; Smith et al., 2003). Recall of a typical day is a semanticised memory because it is a recurring experience (Linton, 1982; Shimamura, 2002). Whether or not this effect applies to the re-telling of uniquely personal (and therefore episodic rather than semanticised) memories in TLE has yet to be studied. Retrieval of autobiographical memories involves a rich recollection of personal events by capturing affectively charged multi-sensory representations of events (Tulving, 1983). As opposed to the highly semanticised retelling of a ‘Typical Day’ (D’Aprano et al., 2022a pre-print), personal reminiscence relies on retrieving a specific event uniquely contextualised in time and space, and depends on the preservation of order and associative structure during formulation (Conway & Pleydell-Pearce, 2000; Tulving, 1983).

This study aimed to understand how individuals with TLE deal linguistically with the retelling of a personal reminiscence at the microlinguistic (lexical-syntactic) and macrolinguistic (suprasentential) levels. We hypothesise that in TLE, compared with healthy controls, retelling a specific, deeply personal memory will be disrupted by macrolinguistic dysfunction, impacting discourse production and refinement. We anticipate this effect to manifest as a failure to eliminate peripheral or redundant details, reflecting a general tendency to produce a vague, non-specific account, and inefficiency in refining the mental representation of a personal story.

## Methods

### Participants

This study included 29 participants, 15 with focal unilateral TLE comprising 10 left TLE (5 mesial, 3 neocortical, 2 non-lesional) and 5 right TLE (4 mesial, 1 neocortical), and 14 healthy controls. Participation required a diagnosis of drug-resistant TLE at recruitment (Kwan et al., 2010), no prior neurosurgical resection, English as a first language, full scale IQ > 70, no reported history of substance-related and addictive disorders, no formally diagnosed psychiatric disorders, and no current major psychiatric episode (e.g., psychosis). None of these individuals were receiving additional treatments for the control of seizures (e.g., vagal nerve stimulation) and none had a history of developmental language disorder, and no neurological condition unrelated to their epilepsy (e.g., stroke). These participants underwent multi-day video electroencephalography (video-EEG) in either The Royal Melbourne Hospital or the Alfred Hospital in Melbourne, Australia to localise seizure focus. In these settings, diagnostic decisions are made by comprehensive teams of neurologists, epileptologists, neurophysiologists, psychiatrists, and neuropsychologists in accordance with International League Against Epilepsy criteria (ILAE; Engel Jr, 2006). Seizure semiology, video-EEG, magnetic resonance imaging (MRI), positron emission tomography (PET) and inter-ictal single-photon emission computed tomography (SPECT) were used to reach unambiguous diagnoses with a localised seizure focus. By assessment, three individuals with TLE reported no seizures in the preceding 12 months on their current anti-seizure medication (ASM) regimen (Kwan et al., 2010). Analyses were performed both with and without these drug-responsive participants to determine whether their inclusion was a robust choice. We found no difference in key outcome measures and these individuals were subsequently retained in the sample. Their reduced epilepsy burden at that point in time is reflected in the 13-point Seizure Frequency Rating (So et al., 1997)—a composite metric capturing seizure frequency, type, and the need for ASM. Family members or partners of participants with TLE were recruited as healthy controls, and where necessary were drawn from the community via convenience sampling to age-, education-, and sex-match to those with TLE. Their demographic characteristics are reported in Table 1 and are broadly comparable on variables of interest.

**Table 1.** Sample characteristics.

*Sample characteristics*
| Variable | Control<br>(*n* = 14) |  | TLE<br>(*n* = 15) |  | *p* | Effect size |
| --- | --- | --- | --- | --- | --- | --- |
|  | Median (*Q1*, *Q3*) | Range | Median (*Q1*, *Q3*) | Range |  |  |
| Age [years] | 51.50 (26.00, 55.50) | 50 | 43.00 (32.00, 53.50) | 44 | 0.96 | 0.01 |
| Education [years] | 15.50 (12.00, 18.00) | 9 | 15.00 (12.00, 17.00) | 15 | 0.58 | 0.12 |
| Estimated IQ<sup>a</sup> | 107.75 (97.63, 111.88) | 41 | 102.50 (95.25, 107.00) | 50.50 | 0.20 | 0.29 |
| Age at Diagnosis [years] | N/A | N/A | 25.00 (20.00, 43.50) | 50 |  |  |
| Epilepsy duration [years] | N/A | N/A | 7.00 (3.00, 17.50) | 41.50 |  |  |
| Seizure Burden Rating | N/A | N/A | 6.00 (2.00, 7.00) | 8 |  |  |
| Last seizure [weeks] | N/A | N/A | 8.00 (4.00, 40.00) | 75 |  |  |
| Current ASMs | N/A | N/A | 2.00 (1.00, 2.00) | 3 |  |  |
| Laterality, Left [n, %] | N/A | N/A | 10 (67) |  |  |  |
| Mesial focus [n, %]<sup>^</sup> | N/A | N/A | 9 (60) |  |  |  |
| Sex, Female [n, %] | 7 (50) |  | 7 (47) |  | 0.86 | 0.03 |
| Handedness, Right [n, %] | 12 (86) |  | 12 (80) |  | 0.68 | 0.08 |

*Note.* TLE = Temporal Lobe Epilepsy; ASM = Anti-seizure Medication. For continuous variables, group differences were computed using Mann-Whitney U test, effect size reported is the rank biserial correlation coefficient. <sup>a</sup>Suggests that data does not violate assumptions of normality on Shapiro-Wilk. For discrete variables, percentages of each group reported, group differences computed using $\chi^2$ test of independence, effect size reported is phi-coefficient. Estimated IQ reflects a composite between TOPF scaled score and WASI-II FSIQ-2 score. Seizure frequency calculated on 13-point rating scale (0-12) considering ASM usage, frequency, and type of seizure (So et al., 1997). <sup>^</sup>Of these individuals 5 had a diagnosis of left TLE. Non-mesial cases comprise 4 neocortical (3 left, 1 right), and 2 non-lesional (left seizure focus).

This multi-site study received ethical approval from the Melbourne Health Human Research Ethics Committee in accordance with ethical standards of the 1964 Declaration of Helsinki. All participants provided written informed consent.

### Neuropsychological Assessment and Elicitation of Personal Discourse

All participants underwent neuropsychological and language assessment conducted by a single clinical neuropsychologist. Given the COVID-19 lockdown conditions in Melbourne, Australia at the time of collection, these assessments were completed via telehealth. Multiple metrics were used to examine lexical retrieval at the single-word level including the Boston Naming Test (BNT) (Kaplan et al., 1983), the Controlled Oral Word Association Test (COWAT) (Strauss et al., 2006), the Auditory Naming Test (ANT) (Hamberger & Seidel, 2003) with minor modifications to suit the Australian lexicon, and the Verb Generation Task (VGT) developed to examine verb retrieval (Appendix A).

To elicit discourse relating to an autobiographical memory, participants were asked to tell a positive, personal, specific memory on four immediately consecutive occasions (see Appendix B for script). Once participants indicated that they had finished telling the story, they were asked to “tell me the same story again now please” for a total of four elicitations. No time limit was imposed, participants were not interrupted for the duration of their output and the researcher minimised verbal and non-verbal participation throughout the elicitations.

### Recording and Transcription

Audio output was recorded from Zoom (Zoom Video Communications Inc, 2016) then manually transcribed verbatim and segmented by a single researcher within four weeks of the file being obtained. Statements were segmented so that a single statement refers to a predicate and its corresponding arguments (Stein & Glen, 1979; Trabasso & van den Broek, 1985). This allows for a precise and consistent proposition-based extraction of content and analysis of coherence (Davis et al., 1997), rather than by communicative unit (C-unit). Audacity^®^ software was used to manually extract pause lengths in grammatical and non-grammatical junctures. Sample lengths refer to the total number of completed words within a sample, excluding words that fill pauses such as “um” “uh” (Nicholas & Brookshire, 1993; Stockbridge et al., 2021).

### Coding for Discourse Variables

A set of discourse variables were selected to examine discourse at both the micro- and macrolinguistic level, see Appendix D for a description of all nodes coding. These variables were drawn from multiple models of discourse production and disturbances in clinical populations to conduct a multi-level discourse analysis (Sherratt, 2007). Using NVivo 12 software, a single reviewer with expertise in cognition and linguistics coded all transcripts and was blind to participant characteristics, other than data acquisition date. Each transcript was coded twice by the same reviewer, with an intra-rater agreement of 94% across both coding occasions. Where there were discrepancies, the researcher re-examined the coding criteria to come to a final decision. If there were ambiguities in the criteria for coding, these were clarified and amended.

#### Coding Agreement

In line with other similar analyses of discourse, a second expert reviewer blindly coded 12.5% of transcripts (Sherratt, 2007), being five total transcripts selected at random. The second reviewer had access to the complete, disambiguated codebook for this process. Inter-rater agreement was determined on a point-by-point basis in terms of the specific node to allocate as well as appropriate statement segmentation. All nodes and the segmentation of statements had 92% agreement—both surpassing the minimal accepted requirement level of 80% (Kazdin, 1982). Coding discrepancies were discussed among the reviewers and resolved by consensus. Once again, any ambiguities in the node descriptions were resolved and their coding was updated (Appendix D).

### Statistical Analyses

We used the *Jamovi* software (Jamovi, 2021) to compute group-specific measures of central tendency, to assess group differences, and to calculate partial correlations. Many of the data were skewed and to be conservative, non-parametric tests (Mann-Whitney U test) have been applied across all analyses for the purpose of uniformity. Data that did not violate assumptions of normality are indicated in table notes. Contingency tables (χ^2^ test of independence) were used for categorical variables. The rank biserial correlation (RBC) was used as a non-parametric estimate of effect size, reflected as small 0.1 < 0.3 < 0.5 to large. Partial correlations were used to examine the relationships between core discourse variables at Trial 5, demographic, and seizure characteristics while adjusting for age. These are reported as Spearman’s rank correlation coefficients.

To ensure analyses were not sensitive to an artifact such as group, we ran the analyses with left and right TLE separately and then together. These analyses showed no difference in discourse outcomes between left and right TLE or relative to controls when individuals with TLE were considered as separate left and right groups or a single group. Consistent with the notion that high-level language functions are not lateralised, all individuals with TLE were subsequently treated as a single group. This methodological point is further addressed in the discussion. To account for Type I error, a false discovery rate (FDR) of 0.05 was applied to primary analyses (Benjamini & Hochberg, 1995). General linear mixed models (GLMMs) were estimated using the *GAMLj* package for the *Jamovi* software, with group as factor, participant as cluster variable, and trial as covariate. Following sensitivity analysis, age was included as an additional covariate for production rate, and seizure burden was included as an additional covariate for sample length, spontaneous duration, pause duration, fluency disruptors, and non-grammatical hesitations.

## Results

### Sample characteristics

Individuals with TLE and controls were comparable across demographic characteristics and many aspects of neuropsychological function (Table 1 and Appendix C, Table 1). Individuals with TLE reported higher rates of depressive symptomatology than controls. While not reflected in total raw or scaled scores for number of correct items on metrics of lexical retrieval, individuals with TLE demonstrated a lexical retrieval deficit when examining delays—with increased mean response time latencies and increased tip-of-the-tongue (TOT) states, i.e., responding >2000ms post-stimulus or requiring phonemic prompting. The same effect was seen for BNT and ANT metrics of total TOT states, TOT states as a proportion of responses, and mean response latencies. Word findings difficulties were reported on Hamberger and Seidel’s (2003) rating scale to be more frequent and more distressing than controls. Individuals with TLE also demonstrated longer latencies on VGT and produced fewer words within a semantic category.

### The TLE phenotype in repeated personal discourse

Group differences on main discourse metrics at each trial are reported in Appendix C, Table 2. The majority of these metrics are calculated relative to sample length or number of total statements. On the initial telling (Trial 1), individuals with TLE were less fluent and cohesive than controls. This manifested as a slower rate of production (words/second), more hesitations, clarity disruptors, and cohesion disruptions, including more personal referents that were ambiguous, incomplete, or missing. Clarity disruptors include instances of circumlocution, deictic terms, empty phrases, and indefinite terms which contribute to the volume of overall output without adding to content or informativeness and disrupt the fluency and overall quality of output. An example from a patient with TLE at Trial 3 exemplifies a combination of these disturbances which are underlined: “<u>yeah like</u> there’s pictures of us eating chicken nuggets and <u>all this different stuff so yeah like</u> I don’t remember it but there’s definitely <u>like</u> proof that we had a proper party <u>and yeah</u> I flew home really sick <u>and then yeah so on and so forth</u>”. Other examples of clarity disruptors commonly identified among individuals with TLE include “sort of thing”, “you know”, “things like that”, “all that sort of stuff”. While these effects on fluency and cohesion broadly persisted across trials, by Trial 2 clear differences in informativeness emerged, compromising coherence in TLE. These comprised fewer novel statements, more statements that did not progress content (non-progression units), and a higher ratio of non-progression to novel units. After four consecutive repetitions, individuals with TLE remained less fluent, less cohesive, and less coherent. The effect sizes for core discourse variables can be visualised in Figure 1.

**Figure 1.**
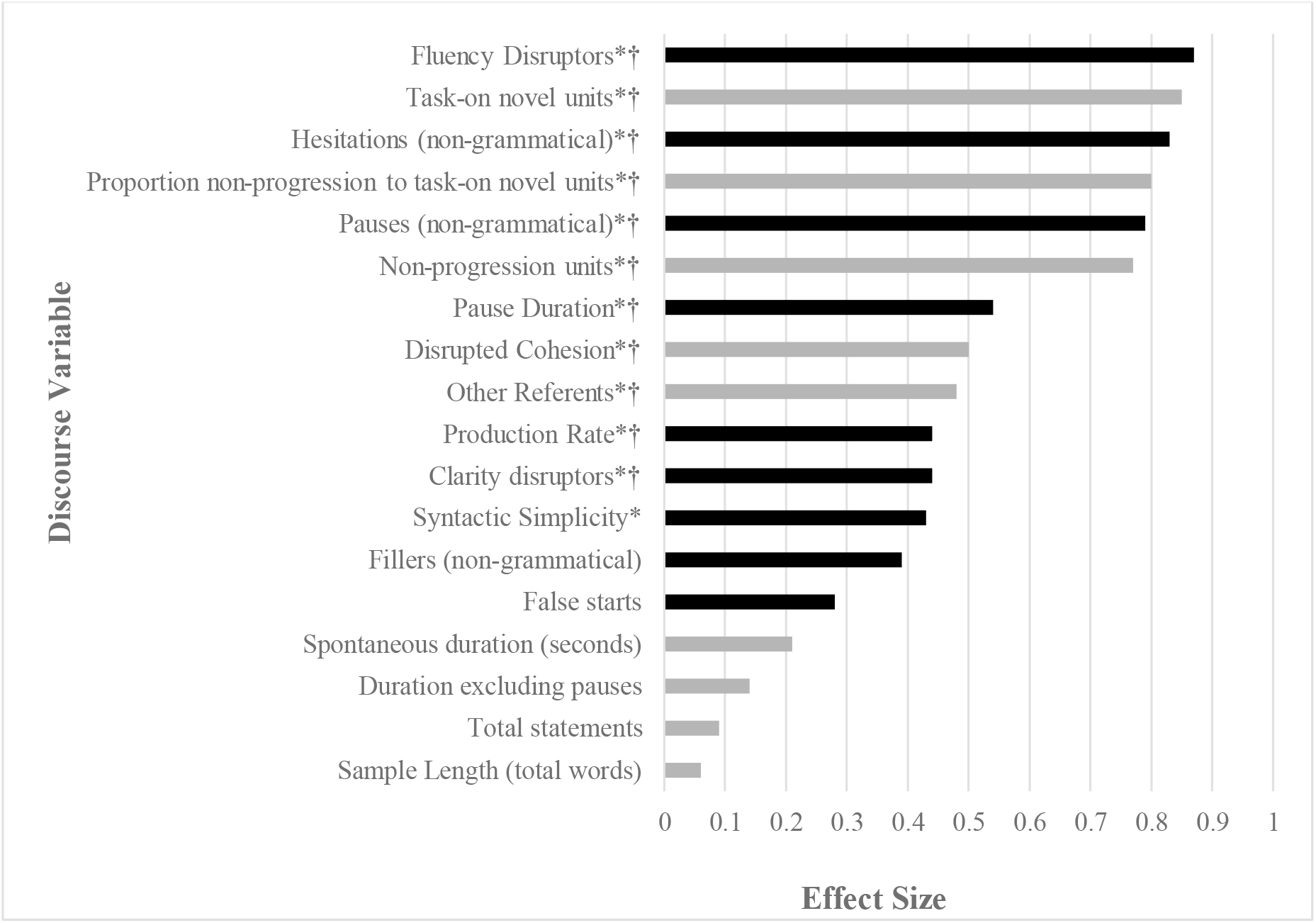
Mean differences on discourse variables between TLE and controls for the autobiographical memory recall task at final trial (Trial 4), represented as absolute value effect size (rank biserial correlation, small 0.1 < 0.3 < 0.5 large), * = *p* < .05, † = significance holds on false discovery rate (FDR) correction. Microlinguistic features are represented in black, macrolinguistic features are grey.

Partial correlations, adjusting for age, were used to examine relationships between clinical characteristics and Trial 4 discourse variables where there were significant group differences: pause duration, production rate, fluency disruption, cohesion disruption, and non-progression to novel units. These are reported in Supplementary Material, Appendix E.

### Change across trials: General Linear Mixed Models

For core metrics of verbosity and informativeness, distinctions between individuals with TLE and controls emerged at Trial 2 and we see a continuation and oftentimes deepening of this effect over additional repetitions. Covarying for age and seizure burden did not impact the fixed effects for total statements, clarity disruptors, non-grammatical pauses, syntactic simplicity, cohesion disruptors, other referents, task-on novel units, non-progression units, or the proportion of non-progression to task-on novel units. As such, the models are reported without these covariates. For production rate there was a significant effect of age; for sample length, spontaneous duration, pause duration, fluency disruptors, and non-grammatical hesitations, there was a significant effect of seizure burden, and the models for these variables reflect their inclusion. Effects plots for models with significant interaction effects can be visualised in Figure 2.

**Figure 2.**
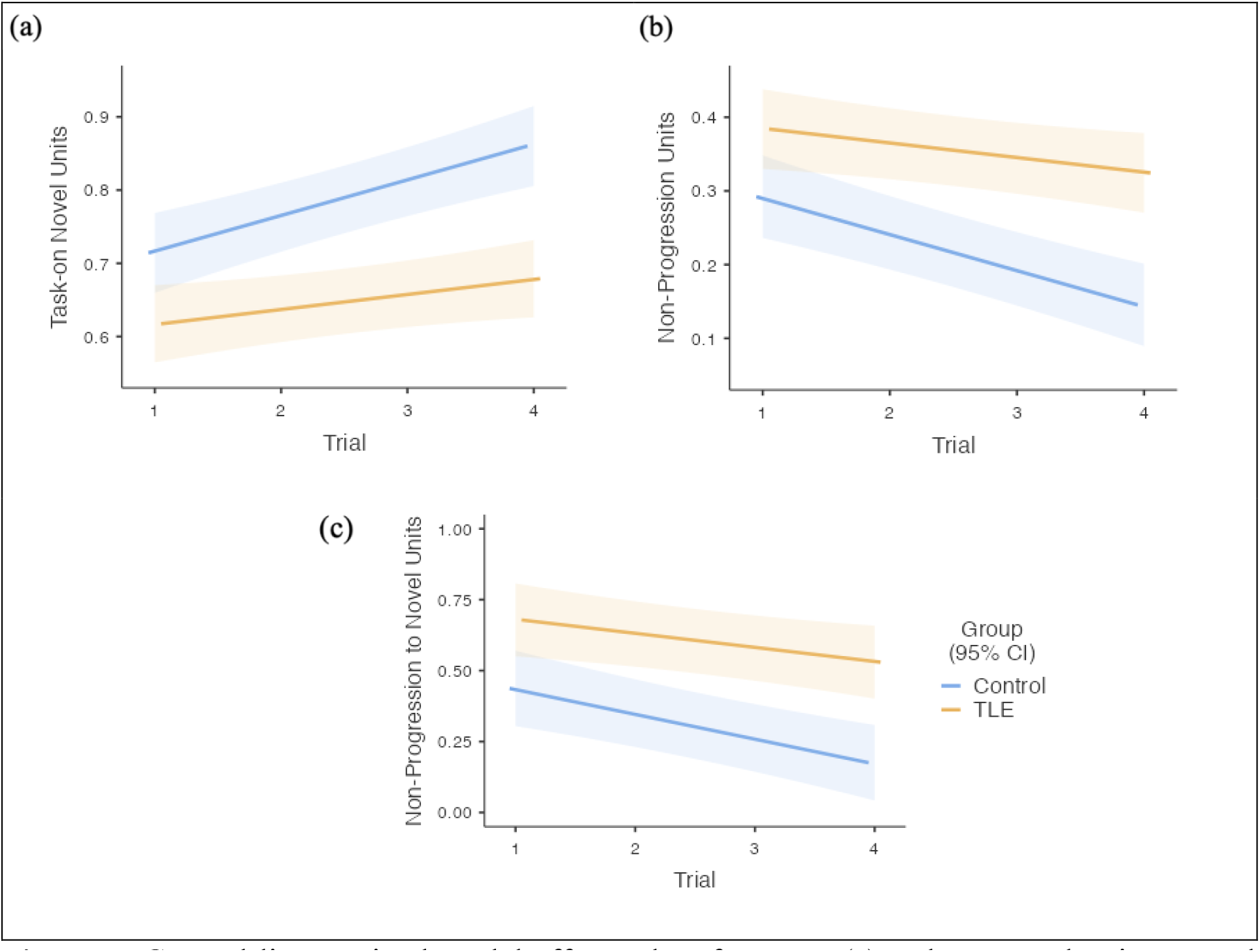
General linear mixed model effects plots for mean (a) task-on novel units to total statements; (b) non-progression units to total statements; (c) proportion of non-progression to task-on novel units across trials in TLE and controls. Trial by group interaction effects are statistically significant. Errors bars represent 95% confidence interval.

#### Output volume

There were no significant effects of trial, group, or interaction effects for sample length, spontaneous duration, or number of total statements.

#### Fluency

For production rate, there was a significant effect of age, *F*(1, 26) = 11.42, *p* = .002. When adjusting for this, both groups significantly increased production rate, with a significant effect of trial, *F*(1, 85) = 9.71, *p* = .0025; and a significant effect of group, *F*(1, 26) = 15.26, *p* = .0006, where controls had a consistently faster production rate than those with TLE. For pause duration, there was a significant effect of trial, *F*(1, 85) = 10.35, *p* = .0018, but no significant effect of group, nor trial by group interaction. For the number of fluency disruptors, there was a significant effect of trial, *F*(1, 85) = 26.82, *p* <.0001; group, *F*(1, 26) = 28.58, *p* < .0001; and seizure burden *F*(1, 26) = 6.12, *p* = .020, but no significant interaction. While the number of fluency disruptors decreased across trials for all participants, individuals with TLE had consistently more disruptors than controls. There was a significant effect of group on the number of clarity disruptors, with TLE producing consistently more than controls, *F*(1, 27) = 13.72, *p* = .0010. There was no significant effect of trial, nor trial by group interaction. For non-grammatical pauses there was a significant effect of trial, *F*(1, 85) = 30.89, *p* < .0001; group, *F*(1, 27) = 27.52, *p* < .0001; but no trial by group interaction. Both groups decreased the number of non-grammatical pauses across trials, although those with TLE produce consistently more than controls. A similar pattern was seen for non-grammatical hesitations with a significant effect of trial, *F*(1, 85) = 18.73, *p* <.0001; group, *F*(1, 27) = 25.88, *p* <.0001; and seizure burden, *F*(1, 27) = 4.54, *p* = .043, and no significant trial by group interaction. There were no significant effects for trial, group, or trial by group interactions for syntactic simplicity.

#### Cohesion

Individuals with TLE produced consistently more cohesion disruptions than controls, as demonstrated by a significant effect of group, *F*(1, 27) = 14.65, *p* = .0007. There is no significant effect of trial nor trial by group interaction. A similar relationship is seen for other referents, being those that are ambiguous, incomplete, or missing, where there is a significant effect of group *F*(1, 27) = 14.15, *p* = .0008, but no significant effect of trial or trial by group interaction.

#### Coherence

There were significant group by trial interaction effects for task-on novel units, non-progression units, and the ratio of non-progression to novel units (see Figure 2). For task-on novel units, there was a significant effect of trial, *F*(1,85) = 23.65, *p* < .0001; group *F*(1, 27) = 19.74, *p* = .0001; and trial by group interaction, *F*(1, 85) = 3.91, *p* = .049. This effect was largely driven by the increase in novel units by controls and stability across trials for those with TLE. The opposite pattern was seen in controls for non-progression units with a progressive decline in the number of non-progression units across trials while there was greater stability in TLE, demonstrated by a significant effect of trial, *F*(1, 85) = 27.24, *p* < .0001; group *F*(1, 27) =16.58, *p* = .0004; and trial by group interaction *F*(1, 85) = 4.89, *p* = .030. When examining percentage change across trials, controls decreased their ratio of non-progression to novel content across the four elicitations by mean 51.42% (*SD* = 38.08), while for TLE this was 20.08% (*SD* = 35.72) and suggestive of greater stability in TLE output. This was demonstrated in the proportion of non-progression to task-on novel units with a significant effect of trial, *F*(1, 85) = 19.91, *p* < .0001; group, *F*(1, 27) = 13.99, *p* = .0009; and trial by group interaction, *F*(1, 85) = 3.97, *p* = 0.049.

## Discussion

This study addresses how individuals with TLE deal linguistically with personal reminiscence and its retelling. Their discourse was marked by disruptions of fluency, clarity, cohesion, and coherence across trials. These disruptions included greater use of hesitations in non-grammatical junctures, ambiguous or missing personal referents, and clarity disruptors in the form of non-specific elements or empty phrases. While improving fluency across repetitions, individuals with TLE had a consistently slower production rate and more fluency disruptions than controls, including pauses, fillers, and false starts. In line with expectations, a pattern of stagnation emerged across repetitions in TLE. Their personalised narratives began and remained vague, producing less novelty and a higher proportion of content that did not progress the story. Extraneous and repetitive details disrupted global coherence and informativeness, similar to the observations of (Marini, 2012). The result was an inefficient communication of personal discourse that became neither more refined nor more detailed across repetitions.

We previously found that issues of increased quantity of output predominated when individuals with TLE told unstructured personal narratives, such as the highly semanticised recount of a typical day (D’Aprano et al., 2022a preprint). In this context, their prolixity was thought to relate to the nature of the elicitation which combines multiple familiar experiences encoded in memory, is less limited in its content and lexicosemantic space, and vulnerable to verbosity (D’Aprano et al., 2022a preprint). These findings stand in contrast to the more nuanced high-level language disturbances we have now observed when personal memories are re-told. For personal reminiscence, discourse production in TLE instead suggested that *quality* rather than quantity, manifesting as inefficient production of a coherent personal account across retellings, was the foreground feature. Repetition is thought to decrease online processing demands associated with initial planning and formulation (Bloom, 1994; Goldman-Eisler, 1968), and is accompanied by increasing recruitment of frontal association cortex (Lillywhite et al., 2010). While retelling benefits discourse processing and refinement for neurologically normal individuals in the form of discourse compression for structured impersonal narratives, such as cartoon images, the same benefit is not seen in TLE (D’Aprano et al., 2022b; Field et al., 2000). As the findings we report here suggest, this also applies to deeply personal discourse in TLE where there is no evidence of compression across trials; that is, no reduction in output volume and duration across repetitions. In overview, the pattern of macrolinguistic impairment we found in TLE likely relates to the nature of the elicitation paradigm and the cognitive-linguistic demands of recounting and reconstructing deeply personal memories.

Reminiscence involves one-off and uniquely personal events contextualised in specific temporal, locational, and emotional frames, placing special demands on cognitive-linguistic systems. By their nature, personal memories rely on broad neural networks beyond those involved in retelling narratives based on pictorial stimuli or semanticised representations. Autobiographical memories are reconstructed each time they are told (Alea & Bluck, 2003; Conway et al., 2004; Kuhlen & Brennan, 2010; Saling et al., 2017). This dynamic process of reconstruction requires individuals to generate and sequence relevant ideas and details of the event, hold these in working memory, self-monitor the evolving narrative, and clarify ambiguities (Alea & Bluck, 2003; Conway et al., 2004; Kuhlen & Brennan, 2010; Reiser et al., 1985). As a result, processing demands do not decrease over time for personal memories, and are not subjected to progressive automatization (Saling et al., 2017). The manner in which individuals with TLE deal linguistically with specific autobiographical memories therefore differs from other unstructured discourse contexts.

At a cognitive level, failure to benefit from retelling in TLE is thought to relate to capacity limitations (Field et al., 2000; Howell et al., 1994; Johns et al., 2008). This impedes how well the mental representation of discourse can be tracked (Bloom, 1994; Mozeiko et al., 2011). Processes related to retrieving specific personal memories place unique challenges on working memory systems (Reiser et al., 1985; Tulving & Thomson, 1973). These processes are critical to pre-planning and organising key event details, holding them in mind, and relating them in an appropriate order to the listener. Our findings suggest that individuals with TLE demonstrated a limited capacity to actively maintain the mental representation of personal discourse, which impacted fluency and cohesion across repetitions. Rather than drawing on novel concepts, capacity limitations in TLE appeared to limit these individuals to concepts they had already retrieved, or alternatively led to the production of more clarity disruptors which are often generic, formulaic, and readily accessible (see Van Lancker Sidtis & Sidtis, 2018 for review). While clarity disruptors can provide support to language planning and formulation by enhancing fluency, there is also a cost. This includes non-specific, imprecise, or extraneous elements which contribute to the volume of output without being informative (Sherratt, 2007). Clarity disruptors, while often fluent in and of themselves, create vague and ambiguous discourse which can impede cohesion and continuity and affect the overall flow of output (Ripich & Terrell, 1988; Sherratt, 2007). They reflect the inability to maintain the discourse representation in mind (Bloom et al., 1993). Clarity disruptors were not prominent in TLE when impersonal structured narratives were retold, where fluency disruptions could presumably be compensated for by shifting to describe a new aspect of the visual stimulus and there are minimal demands on working memory (D’Aprano et al., 2022b preprint).

Pragmatic disturbances impact the capacity to tailor and modify language to suit the conversational context and reflect a communication disorder (Alea & Bluck, 2003; Bluck, Alea, Baron-Lee, & Davis, 2016; Broeders, Geurts, & Jennekens-Schinkel, 2010). Atypical social cognition, including Theory of Mind, is recognised as an accompanying feature of TLE (Broicher et al., 2012; Giovagnoli et al., 2011, 2013; Schacher et al., 2006), and many of these patients experience a lifetime of social dislocation (Wilson et al., 2007). These disturbances emerge in the discourse of individuals with TLE. They appear to be impeded in their capacity to determine what is and is not relevant to an interaction. Telling and retelling deeply personal memories is highly socially determined (Pasupathi & Rich, 2005) and personal reminiscence is non-routinised and cannot be semanticised. The intention is not only to communicate what is relevant, but also what is entertaining and face-saving and might serve to compensate for lower levels of novelty in discourse (Field et al., 2000). Story asides provide opportunities to communicate additional meaning and are more relevant for autobiographical memory recall than impersonal stories (Bluck et al., 2016), where adding extraneous details or becoming repetitive likely has a stronger social-motivation component (Broicher et al., 2012; Schacher et al., 2006). Unlike controls who can maintain interest by introducing novelty, disturbances to the depth of detail and temporal sequencing of events in TLE (Addis et al., 2007; Rosenbaum et al., 2005; St-Laurent et al., 2011; Steinvorth et al., 2005; Vargha-Khadem et al., 1997; Viskontas et al., 2000) might preclude them from doing so as efficiently. Their compensation might instead be limited to statements that are repetitive, extraneous, or non-specific and which disrupt clarity. This is consistent with clinical observations of a ‘sticky’ interpersonal style (Rao et al., 1992).

While more fundamental aspects of language function can be lateralised, this is not the expectation for higher-order language which is more likely to be represented in tertiary cortex and therefore less likely lateralised (Dick & Tremblay, 2012; Johns et al., 2008; Schneider et al., 2021; Xu et al., 2005). In light of this, right and left TLE participants have been considered as a single diagnostic group. This approach was supported by our analyses which indicated that there were no differences in discourse outcomes relative to controls when participants with TLE were considered as separate left and right groups or collectively, and ultimately that treating them as a single group was a robust choice. The sample size for this study, while small in general research design terms, was exhaustive for Victorian hospitals at the time of collection. The detailed linguistic analysis generated a very large corpus of data points for each participant. Given the social determinants of discourse, the interaction with the researcher should be considered for its potential influence on language samples. Feedback from the listener and their attentiveness is known to influence how a narrative is produced (Kuhlen & Brennan, 2010; Pasupathi et al., 1998). Researcher characteristics and facial expressions might play a role in the discourse produced based on their age, gender, ethnicity, level of interest expressed, and overall rapport. The researcher aimed to provide consistently warm and attentive reactions to the description of all stories, and had their microphone muted to avoid verbal contributions. The same researcher conducted all data collection and therefore interpersonal factors were more likely to be consistent throughout.

The study is the first to examine how individuals with TLE deal linguistically with personal narrative. The macrolinguistic phenomenon we have identified is influenced by the elicitation paradigm. For personal reminiscence, there is a disturbance in the quality rather than the quantity of output, where content familiarity might conflict with the diverse cognitive-linguistic demands of planning and producing spontaneous personal discourse.

## Data Availability

All data produced in the present study are available upon reasonable request to the authors

