## Appendix B for "Vague retellings of personal narratives in temporal lobe epilepsy"

### Autobiographical Memory Recall Script

“I’d like you to take a few minutes now to think about a happy memory from your more recent life, something from more than a year ago but within the last ten years. It can be anything, as long as the event was positive for you and that you are able to retell it to me in detail. Do not pick an event that you heard about from others. It must be from a specific time and place, and something you were personally involved in. For example, playing basketball on Thursday nights would not be specific enough. However, you could tell me about one specific game where you fell, broke your arm, and needed to go to the hospital etc. I want you to tell me as much detail as you can about the event. (120 seconds to lapse). You may begin telling me the story and when you think you have told me everything you can, let me know”.
