## Appendix C for "Vague retellings of personal narratives in temporal lobe epilepsy"

**Table 1**

*Sample characteristics for neuropsychological variables*

| Variable | Control (*n* =14) Median *(Q1,Q3)* | Control (*n* =14) Range | TLE (*n* =15) Median *(Q1,Q3)* | TLE (*n* =15) Range | *p* | *r* |
|---|---|---|---|---|---|---|
| TOPF[a] | 0.75 (-0.02, 1.33) | 2.25 | 0.25 (-0.67, 0.67) | 3.00 | 0.06 | 0.41 |
| WASI-II $_{\text{Matrix Reasoning}}$ | 0.40 (0.00, 0.92) | 6.50 | 0.00 (-0.33, 0.55) | 4.50 | 0.40 | 0.19 |
| WASI-II $_{\text{Vocabulary}}$ | 1.00 (-0.18, 1.25) | 3.50 | 0.55 (0.14, 0.67) | 3.67 | 0.20 | 0.29 |
| WASI-II $_{\text{FSIQ}}$ | 0.61 (-0.25, 0.75) | 3.17 | 0.25 (-0.42, 0.55) | 4.50 | 0.26 | 0.25 |
| WAIS-IV Digits Forward[a] | 0.34 (-0.67, 0.92) | 3.67 | -0.33 (-0.33, 0.17) | 2.33 | 0.47 | 0.16 |
| WAIS-IV Digits Backward[a] | 0.00 (-0.67, 0.33) | 3.34 | 0.00 (-0.33, 0.50) | 2.66 | 0.98 | 0.01 |
| WMS-IV Logical Memory I[a] | 0.67 (-0.16, 1.33) | 4.00 | 0.33 (-0.17, 1.00) | 2.33 | 0.58 | 0.12 |
| WMS-IV Logical Memory II[a] | 1.00 (0.00, 1.00) | 3.66 | 0.33 (-0.33, 1.00) | 2.67 | 0.48 | 0.16 |
| RAVLT $_{\text{Total}}$[a] | 0.59 (-0.09, 1.52) | 3.57 | 0.18 (-0.15, 1.15) | 3.01 | 0.62 | 0.11 |
| RAVLT $_{\text{Post-Interference Recall}}$[a] | 0.22 (-0.58, 1.10) | 3.01 | 0.22 (-0.86, 0.89) | 3.76 | 0.65 | 0.10 |
| RAVLT $_{\text{Delayed Recall}}$[a] | 0.28 (-0.46, 1.26) | 4.34 | 0.32 (-0.77, 0.69) | 3.91 | 0.65 | 0.10 |
| WMS-R Easy^ | 12.00 (11.00, 12.00) | 3.00 | 11.00 (10.00, 11.50) | 3.00 | 0.18 | 0.28 |
| WMS-R Hard^[a] | 8.50 (6.25, 10.00) | 10.00 | 7.00 (6.00, 8.50) | 9.00 | 0.21 | 0.28 |
| WMS-R Easy Delay^ | 4.00 (4.00, 4.00) | 0.00 | 4.00 (4.00, 4.00) | 1.00 | 0.37 | 0.07 |
| WMS-R Hard Delay^ | 4.00 (3.25, 4.00) | 4.00 | 4.00 (3.00, 4.00) | 2.00 | 0.60 | 0.10 |
| Victoria Stroop $_{\text{Dots}}$[a] | -0.83 (-1.25, -0.33) | 2.33 | -1.33 (-1.33, -1.00) | 3.34 | 0.12 | 0.34 |
| Victoria Stroop $_{\text{Words}}$[a] | -0.50 (-0.92, -0.33) | 1.67 | -0.67 (-1.33, -0.33) | 2.33 | 0.31 | 0.22 |
| Victoria Stroop $_{\text{Colour}}$[a] | 0.00 (-0.92, 0.67) | 4.00 | 0.00 (-0.67, 0.33) | 2.00 | 0.60 | 0.12 |
| Victoria Stoop $_{\text{Interference}}$[a] | 0.83 (-0.83, 1.00) | 3.33 | 1.00 (0.50, 1.33) | 2.00 | 0.41 | 0.18 |
| BNT $_{\text{Total}}$[a] | -0.05 (-0.67, 0.55) | 3.03 | -0.74 (-1.65, 0.13) | 3.84 | 0.12 | 0.35 |
| BNT $_{\text{TOT States}}$^[a] | 17.00 (13.75, 23.50) | 22.00 | 24.00 (21.00, 27.00) | 17.00 | 0.02* | 0.48 |
| BNT $_{\text{Proportion TOT}}$^[a] | 0.28 (0.23, 0.40) | 0.37 | 0.40 (0.35, 0.44) | 0.20 | 0.08 | 0.39 |
| OLR (COWAT)[a] | -0.53 (-0.98, -0.04) | 4.01 | -1.17 (-1.58, 0.16) | 4.29 | 0.29 | 0.23 |
| Animals[a] | 0.67 (-0.02, 1.63) | 3.34 | -0.35 (-0.61, 0.30) | 2.39 | 0.0099** | 0.57 |
| ANT $_{\text{Total}}$^ | 48.00 (48.00, 49.00) | 5.00 | 47.00 (45.00, 49.00) | 9.00 | 0.29 | 0.23 |
| ANT $_{\text{Latency}}$^[a] | 64.00 (59.50, 69.00) | 23.00 | 84.00 (68.50, 92.00) | 60.00 | 0.02* | 0.53 |
| ANT $_{\text{TOT States}}$^[a] | 11.50 (9.00, 14.50) | 18.00 | 21.00 (13.00, 27.00) | 29.00 | 0.03* | 0.47 |
| ANT $_{\text{Proportion TOT}}$^[a] | 0.23 (0.18, 0.29) | 0.36 | 0.42 (0.26, 0.54) | 0.58 | 0.03* | 0.47 |
| WFD Frequency Rating^[a] | 2.50 (1.25, 3.00) | 3.00 | 3.00 (3.00, 4.00) | 5.00 | 0.04* | 0.45 |
| WFD Distress Rating^ | 1.00 (1.00, 1.75) | 3.00 | 4.00 (3.00, 6.00) | 6.00 | 0.0006** | 0.74 |
| VGT $_{\text{Correct Dominant}}$^ | 36.50 (34.25, 42.75) | 20.00 | 36.00 (32.00, 38.00) | 22.00 | 0.34 | 0.21 |
| VGT $_{\text{Correct Other}}$^ | 18.00 (15.00, 21.00) | 15.00 | 19.00 (17.00, 21.25) | 14.00 | 0.46 | 0.17 |
| VGT $_{\text{Latency}}$^ | 184.50 (164.25, 196.75) | 125.00 | 213.00 (183.50, 308.00) | 323.00 | 0.03* | 0.48 |
| HADS $_{\text{Anxiety}}$^[a] | 7.00 (5.25, 8.50) | 10.00 | 7.00 (6.50, 8.50) | 10.00 | 0.58 | 0.12 |
| HADS $_{\text{Depression}}$^ | 2.00 (1.00, 3.00) | 9.00 | 4.00 (4.00, 5.50) | 16.00 | 0.022* | 0.50 |

*Note.* TLE = Temporal Lobe Epilepsy; TOPF = Test of Premorbid Functioning; WASI-II = Wechsler Abbreviated Scale of Intelligence, Second Edition; FSIQ = Full Scale Intelligence Quotient; WAIS-IV = Wechsler Adult Intelligence Scale, Fourth Edition; WMS-IV = Wechsler Memory Scale, Fourth Edition; RAVLT = Rey Auditory Verbal Learning Test; WMS-R = Wechsler Memory Scale–Revised; BNT = Boston Naming Test; TOT = Tip-of-the-tongue; OLR (COWAT) = Orthographic Lexical Retrieval (Controlled Oral Word Association Test); WFD = Word Finding Difficulty; VGT = Verb Generation Task; ANT = Auditory Naming Task; HADS = Hospital Anxiety and Depression Scale. Represented as z-scores where normative data was available, raw data are indicated by^. Latencies are expressed in seconds. Group differences computed using Mann-Whitney U test, effect sizes *r*, where * = p <.05, ** = p <.01. [a]Suggests that data does not violate assumptions of normality on Shapiro-Wilk.

**Table 2**

*Group differences on discourse variables across trials*

| Discourse Measure | Trial 1 | | Trial 2 | | Trial 3 | | Trial 4 | |
|---|---|---|---|---|---|---|---|---|
| | *MD* [95% CI] | *p* [RBC] | *MD* [95% CI] | *p* [RBC] | *MD* [95% CI] | *p* [RBC] | *MD* [95% CI] | *p* [RBC] |
| Sample length | 26.50 [-179.00, 300.00] | .71 [0.09] | -12.00 [-228.00, 147.00] | .85 [0.05] | -26.00 [-274.00, 145.00] | .68 [0.10] | -15.74 [-259.00, 154.00] | .79 [0.06] |
| Spontaneous duration | -12.84 [-112.95, 70.83] | .85 [0.05] | -36.05 [-125.79, 35.54] | .38 [0.20] | -37.76 [-132.72, 26.94] | .19 [0.30] | -31.50 [-123.70, 39.90] | .35 [0.21] |
| Pause duration | -9.74 [-30.70, 4.34] | .31 [0.23] | -18.53 [-37.50, -1.71] | .026 [0.49]* | -20.50 [-39.10, -4.78] | .0094 [0.57]*† | -20.38 [-42.36, -1.99] | .008 [0.54]*† |
| Duration (excluding pauses) | -3.96 [-83.87, 65.27] | .92 [0.03] | -14.65 [-86.48, 34.77] | .51 [0.15] | -17.74 [-96.18, 31.88] | .45 [0.17] | -14.06 [-81.09, 44.75] | .53 [0.14] |
| Production rate (words/second) | 0.60 [0.19, 0.99] | .0043 [0.61]*† | 0.57 [0.17, 0.90] | .0043 [0.61]*† | 0.53 [0.10, 0.89] | .014 [0.53]*† | 0.46 [0.02, 0.80] | .032 [0.44]*† |
| Total statements | 5.82 [-24.00, 41.00] | .66 [0.10] | -2.70 [-23.00, 22.00] | .71 [0.09] | -4.18 [-39.00, 19.00] | .57 [0.13] | -3.00 [-37.00, 21.00] | .71 [0.09] |
| Fluency disruptors^ | -0.11 [-0.17, -0.05] | .0006 [0.71]*† | -0.12 [-0.18, -0.07] | <.0001 [0.80]*† | -0.11 [-0.16, -0.76] | <.0001 [0.86]*† | -0.10 [-0.15, -0.06] | <.0001 [0.87]*† |
| Clarity disruptors^ | -0.01 [-0.03, -0.00] | .0068 [0.58]*† | -0.02 [-0.03, -0.01] | .0032 [0.63]*† | -0.01 [-0.02, -0.00] | .0011 [0.69]*† | -0.01 [-0.03, 0.00] | .033 [0.44]*† |
| False starts^ | -0.01 [-0.03, 0.01] | .29 [0.24] | -0.01 [-0.03, 0.00] | .057 [0.42] | -0.01 [-0.03, -0.00] | .033 [0.47]*† | -0.01 [-0.02, 0.01] | .22 [0.28] |
| Fillers (non-grammatical)^ | -0.01 [-0.02, 0.00] | .15 [0.32] | -0.01 [-0.03, 0.00] | .057 [0.42] | -0.01 [-0.03, 0.00] | .016 [0.31]*† | -0.01 [-0.03, 0.00] | .077 [0.39] |
| Pauses (non-grammatical)^ | -0.05 [-0.08, -0.02] | .0019 [0.66]*† | -0.05 [-0.07, -0.02] | .0001 [0.78]*† | -0.04 [-0.06, -0.03] | <.0001 [0.82]*† | -0.05 [-0.07, -0.03] | .0001 [0.79]*† |
| Hesitations (non-grammatical)^ | -0.06 [-0.11, -0.03] | .0008 [0.70]*† | -0.07 [-0.10, -0.03] | .0001 [0.78]*† | -0.06 [-0.08, -0.03] | .0006 [0.75]*† | -0.06 [-0.09, -0.04] | <.0001 [0.83]*† |
| Other referents^ | -0.02 [-0.02, -0.01] | .0003 [0.74]*† | -0.01 [-0.03, 0.00] | .055 [0.42]* | -0.02 [-0.03, -0.01] | .0009 [0.70]*† | -0.01 [-0.02, -0.00] | .019 [0.48]*† |
| Disrupted Cohesion^ | -0.02 [-0.03, -0.01] | .0008 [0.70]*† | -0.01 [-0.03, -0.00] | .040 [0.45]* | -0.02 [-0.03, -0.01] | .0005 [0.72]*† | -0.01 [-0.03, -0.00] | .016 [0.50]*† |
| Task-on novel units# | 0.10 [0.01, 0.18] | .034 [0.47]* | 0.15 [0.04, 0.27] | .0007 [0.58]*† | 0.15 [0.06, 0.26] | .0006 [0.61]*† | 0.19 [0.09, 0.25] | .0001 [0.85]*† |
| Non-progression units# | -0.08 [-0.15, 0.01] | .081 [0.39] | -0.15 [-0.27, -0.04] | .0008 [0.57]*† | -0.16 [-0.28, -0.08] | .0021 [0.68]*† | -0.16 [-0.23, -0.07] | .0005 [0.77]*† |
| Non-progression to task-on novel units | -0.21 [-0.38, -0.00] | .045 [0.44]* | -0.29 [-0.61, -0.06] | .0008 [0.57]*† | -0.27 [-0.57, -0.13] | .00028 [0.66]*† | -0.28 [-0.44, -0.12] | .0003 [0.80]*† |
| Syntactic Simplicity^ | -0.02 [-0.04, 0.00] | .057 [0.42] | -0.02 [-0.04, -0.00] | .029 [0.48]* | -0.02 [-0.04, -0.00] | .029 [0.48]*† | -0.02 [-0.04, -0.00] | .038 [0.43]* |

*Note.* TLE = Temporal Lobe Epilepsy; MD = Mean Difference where Control – TLE, CI = Confidence Interval. Metrics marked ^ are expressed relative to sample length, # are expressed relative to total statements. Group differences computed using Mann-Whitney U test, effect sizes expressed as Rank Biserial Correlation, where * = $p$ <.05, † = significance holds on false discovery rate (FDR) correction.
