## Appendix E for "Vague retellings of personal narratives in temporal lobe epilepsy"

*Partial Correlations*

Partial correlations, adjusting for age, were used to examine relationships between clinical characteristics and Trial 4 discourse variables where there were significant group differences: pause duration, production rate, fluency disruption, cohesion disruption, and non-progression to novel units. Seizure burden correlated significantly with disease duration ($r$ = 0.89, $p$ < .0001), number of ASMs ($r$ = 0.83, $p$ < .0001), time since last seizure ($r$ = 0.78, $p$ < .0001), production rate ($r$ = -0.37, $p$ = .037), pause duration ($r$ = 0.54, $p$ = .003), disrupted cohesion ($r$ = 0.44, $p$ = .018), fluency disruptors ($r$ = 0.49, $p$ = .008), and proportion non-progression to novel units ($r$ = 0.58, $p$ = .0011). Disease duration correlated with age at diagnosis ($r$ = -0.91, $p$ < .0001), number of ASMs ($r$ = 0.90, $p$ < .0001), time since last seizure (r = 0.78, p < .0001), pause duration ($r$ =0.49, $p$ = .009), production rate ($r$ = -0.48, $p$ = .009), fluency disruption ($r$ = 0.55, $p$ = .003), cohesion disruption ($r$ = 0.43, $p$ = .021), and proportion non-progression to novel units ($r$ = 0.58, $p$ = .0012). Number of ASMs correlated with production rate ($r$ = -0.54, $p$ = .0035), fluency disruptors ($r$ = 0.55, $p$ = .003), cohesion disruptors ($r$ = 0.42, $p$ = .027), and proportion non-progression to novel units ($r$ = 0.60, $p$ = .001). Time since last seizure correlated with pause duration ($r$ = 0.38, $p$ = .043), fluency disruptors ($r$ = 0.59, $p$ = .0009), and proportion non-progression to novel units ($r$ = 0.73, $p$ < .0001). Years of education correlated with estimated IQ ($r$ = 0.74, $p$ < .0001). Production rate correlated with fluency disruption ($r$ = -0.59, $p$ = .0010), while pause duration correlated with proportion of non-progression to novel units ($r$ = 0.40, $p$ = .035). Fluency disruptors also correlated with the proportion of non-progression to novel units ($r$ = 0.44, $p$ = .020). For these core variables, there were no other significant associations with demographic or seizure characteristics at Trial 4 when adjusting for age.
